# Y-FIT Ghana: A Study Protocol for a Youth-Led Participatory Approach to Promote Youth-Friendly HIV Self-Testing, Oral and Long-Acting Injectable Pre-Exposure Prophylaxis Uptake in Ghana

**DOI:** 10.64898/2026.02.23.26346606

**Authors:** Gloria Aidoo-Frimpong, Abass Tando Abubakar, Yaa Adutwumwaa Obeng, Winfred Kofi Mensa, Daniel Selase Anyidoho, Ernest Ortsin, Stephen Ayisi Addo, Anthony Affum Awuah, Ucheoma Nwaozuru, Naa Ashiley Vanderpuye, Paul Sowah, Temitope Ojo, Juliet Iwelunmor, YFIT GH Study Team

## Abstract

**Introduction:** Despite progress in HIV prevention, adolescents and young adults in sub-Saharan Africa remain disproportionately affected by HIV. In Ghana, youth aged 15–24 years face persistent barriers to accessing HIV prevention services, including stigma, limited awareness of prevention options, and insufficient availability of youth-friendly services. Emerging and existing prevention approaches, including HIV self-testing, oral, and long-acting injectable pre-exposure prophylaxis offer important opportunities to reduce HIV infections among youth. However, successful uptake depends on delivery strategies that reflect young people’s priorities and lived experiences. The Youth-Friendly Innovation Tools for HIV Prevention in Ghana (Y-FIT Ghana) study uses a youth-led, participatory approach to co-create strategies that support youth-friendly HIV self-testing and PrEP uptake.

**Methods:** Y-FIT Ghana is a mixed-methods, participatory study grounded in Youth Participatory Action Research and Human-Centered Design, and guided by Social Cognitive Theory. The study will follow a three-phase innovation pipeline: (1) a national open call to solicit youth-generated ideas related to HIV prevention; (2) an HIV Innovation Sprint to refine and develop selected concepts; and (3) a one-week bootcamp focused on capacity building and implementation readiness. Adolescents and young adults aged 15–24 years will be recruited nationwide and engaged as co-designers throughout the study. Data collection will include participatory activities, qualitative assessments, and implementation-focused measures to examine feasibility, acceptability, perceived appropriateness, and youth engagement related to HIV self-testing, oral PrEP, and LAI-PrEP. Qualitative data will be analyzed using thematic analysis, and quantitative data will be summarized using descriptive statistics, with integration across methods to inform refinement of youth-friendly HIV prevention strategies.

**Expected Results:** The study is expected to demonstrate that participatory innovation can produce feasible, youth-driven HIV prevention solutions. Participants are anticipated to gain knowledge, leadership skills, and self-efficacy while developing innovative prototypes suited for youth-friendly delivery.

**Ethics and dissemination:** Ethical approval has been obtained from the University at Buffalo (*IRB ID: STUDY00009385*) and the Ghana Health Service (*ID NO: GHS-ERC-030/05/25*) Institutional Review Boards. Findings will be disseminated through peer-reviewed publications, youth- and community-facing outputs, stakeholder engagement activities, and policy-relevant briefs to inform youth-centered HIV prevention efforts in Ghana and similar settings.

## Introduction

Young people across sub-Saharan Africa continue to shoulder a disproportionate burden of the HIV epidemic (UNAIDS, 2024). Despite sustained global progress in HIV prevention, the region accounted for nearly half of all new HIV infections in 2023 (UNAIDS, 2024). Structural and socio-cultural factors, including stigma, limited access to healthcare, and low awareness of prevention options, continue to undermine effective HIV prevention and treatment efforts (Makusha & Gittings, 2024). Within this regional context, adolescents and young adults (AYAs) remain among the most affected and least reached populations. In 2023, approximately 360,000 young people aged 15–24 years were newly infected with HIV worldwide, with adolescents aged 15–19 years accounting for nearly 38.9% of these infections (UNICEF, 2023). Sub-Saharan Africa bears the overwhelming share of this burden, accounting for nearly 84% of the world’s young people affected (UNICEF, 2023). These trends highlight the persistent vulnerability of youth and underscore the need for prevention strategies that are developmentally appropriate, socially responsive, and aligned with young people’s lived realities.

Ghana reflects these broader regional disparities. According to the Ghana AIDS Commission, there were an estimated 17,774 new HIV infections and 12,480 AIDS-related deaths in 2023, with approximately 334,095 people living with HIV nationwide (Ghana Aids Commission, 2024). Although youth aged 15–24 years make up a smaller share of the population, they accounted for nearly 29% of new HIV infections, highlighting a disproportionate burden in this age group. This pattern is shaped by interconnected structural and social constraints that limit young people’s engagement with HIV prevention. Restricted access to youth-friendly services, economic dependence, and limited autonomy in navigating health systems reduce opportunities for confidential testing and prevention, while stigma and fear of social consequences further discourage service use (Castor et al., 2025;Njau et al., 2019). Beyond access barriers, HIV prevention efforts often fail to meaningfully involve young people in shaping how services are designed, communicated, and delivered, leading to approaches that feel misaligned with youth social realities. Consistent with these constraints, only about 25%-45% of Ghanaian youth have ever tested for HIV (Andoh-robertson, 2018; Asare et al., 2020; Eshun et al., 2023), and many remain unaware of available biomedical prevention tools or uncertain about how to access them (Eshun et al., 2023). These intersecting barriers illustrate the urgent need for strategies that align with AYAs’ lived realities and provide more acceptable, accessible, and empowering pathways to HIV testing and prevention. Within this context, prevention strategies that reduce reliance on facility-based services and increase privacy, autonomy, and youth control are especially critical for improving engagement along the HIV prevention continuum.

HIV self-testing (HIVST) has emerged as a promising strategy to address these challenges by shifting HIV testing away from clinical settings and into spaces where young people may feel safer and more in control. By allowing individuals to collect specimens and interpret results privately, HIVST directly responds to concerns related to stigma, confidentiality, and limited autonomy that often deter adolescents and young adults from seeking facility-based testing (Lindquist-Grantz & Abraczinskas, 2018; Nagai et al., 2021; Obeagu & Obeagu, 2024). Evidence from multiple African settings demonstrates that HIVST can increase testing uptake, support earlier diagnosis, and facilitate linkage to prevention services among youth (Iwelunmor et al., 2020; van Empel et al., 2022). As a result, both the World Health Organization and Ghana’s national HIV guidelines recommend HIVST as a complementary testing strategy. Features such as privacy, convenience, and autonomy make HIVST particularly appealing to adolescents and young adults (Nagai et al., 2021). However, despite national policy endorsement, awareness and use of HIVST in Ghana remain limited, and few initiatives have meaningfully engaged youth in shaping how self-testing is promoted or accessed. Without deliberate youth engagement in the design of messaging, access points, and support systems, the potential of HIVST to normalize testing and strengthen linkage to prevention services is unlikely to be fully realized.

Beyond testing, biomedical prevention options such as oral pre-exposure prophylaxis (PrEP) and long-acting injectable PrEP (LAI-PrEP) offer additional opportunities for young people to protect themselves from HIV. Oral PrEP provides effective protection when taken consistently, while LAI-PrEP, administered every two or six months, reduces the need for daily pill-taking and may better align with young people’s preferences for privacy and convenience (Celum et al., 2019; Medina-Marino et al., 2022; Rousseau et al., 2021). Emerging evidence from sub-Saharan Africa suggests that adolescents and young adults are willing to consider LAI-PrEP when information is presented in ways that are relevant, clear, and trustworthy (Kakande et al., 2023; Lunkuse et al., 2025; Schwarzkopf, 2024). In Ghana, however, awareness of PrEP remains low (Deynu & Ouner, 2025). A recent study among educated youth in the Lower Manya Krobo Municipality found that only a small number of participants were aware of PrEP prior to the study, yet brief educational exposure significantly increased interest in its use (Owusu et al., 2024). These findings suggest that limited uptake reflects gaps in information and engagement rather than resistance to biomedical prevention. When combined with HIV self-testing, oral PrEP and LAI-PrEP expand the range of prevention options available to youth, offering greater flexibility and agency across the HIV prevention continuum. Bridging the gap between availability and use requires approaches that place young people at the center of prevention efforts and actively involve them in shaping solutions.

In response to these unmet needs, the Youth-Friendly Innovation Tools for HIV Prevention in Ghana (Y-FIT Ghana) study adapts and extends a social innovation model previously implemented in Nigeria and other low-and-middle-income countries (Idigbe et al., 2023; Iwelunmor et al., 2020; Tan et al., 2024). In Ghana, Y-FIT Ghana establishes a structured social innovation pipeline that positions adolescents and young adults as central actors in the design, refinement, and implementation of strategies to expand HIV self-testing and strengthen demand for oral and long-acting injectable PrEP. Evidence from the Nigerian experience demonstrates that youth-led open calls, design sprints, and mentored bootcamps can generate creative, contextually grounded prevention strategies and can be integrated into implementation systems at scale (Iwelunmor et al., 2020; Iwelunmor, Ezechi, et al., 2022; Iwelunmor, Tucker, et al., 2022; Tan et al., 2024). Building on these insights while tailoring the approach to Ghana’s sociocultural context, Y-FIT Ghana aims to cultivate a national cohort of youth innovators and support them in translating lived experience into actionable prevention strategies. Through this process, the study seeks to generate evidence on the feasibility, acceptability, and early outcomes of a youth-driven social innovation model for HIV prevention in Ghana, with implications for broader application in West Africa.

### Conceptual Framework

Y-FIT Ghana is grounded in Social Innovation Theory, which emphasizes collective intelligence, co-creation, and iterative problem-solving as mechanisms for generating contextually relevant solutions (Bandura, 1986; Bandura & Walters, 1977; Mumford et al., 2002). Within this framework, communities are understood to hold valuable expertise that can be mobilized through inclusive platforms that encourage broad participation. In Y-FIT Ghana, the national open call creates space for diverse youth perspectives, the innovation sprint supports collaborative refinement through peer engagement and mentorship, and the bootcamp provides structured opportunities for capacity building and prototype development. Together, these components foster a youth-driven ecosystem in which prevention strategies emerge from lived experience and are refined through iterative feedback, increasing their relevance, acceptability, and feasibility.

Youth Participatory Action Research (YPAR) provides the participatory foundation for the study by positioning adolescents and young adults as active contributors to knowledge generation and solution development rather than passive recipients of intervention (Fortin et al., 2022; Mathikithela & Wood, 2021; Ozer, 2016). Rather than treating youth as passive beneficiaries, YPAR emphasizes shared ownership, agency, and collective inquiry. Through this lens, young people identify barriers to HIV self-testing and long-acting PrEP uptake, articulate priorities informed by their lived experiences, and co-create strategies that reflect their cultural and social contexts (Iwelunmor, Tucker, et al., 2022). This approach is particularly relevant for HIV prevention, where young people often navigate stigma, confidentiality concerns, and power asymmetries within health systems. By engaging youth as co-researchers, YPAR promotes empowerment, strengthens analytic capacity, and builds trust in the intervention process, increasing the likelihood that resulting innovations will resonate with peers and support sustainable implementation(Anyon et al., 2018; Bennin et al., 2024; Iwelunmor et al., 2020).

Together, Social Innovation Theory and YPAR (Figure 1) enable Y-FIT Ghana not only to generate creative youth-led prevention strategies but also to systematically examine how and under what conditions such innovations can be implemented. This integrated framework lays the groundwork for future scale-up and contributes to the growing evidence base on youth-centered, participatory approaches to HIV prevention in sub-Saharan Africa.

**Figure 1:**
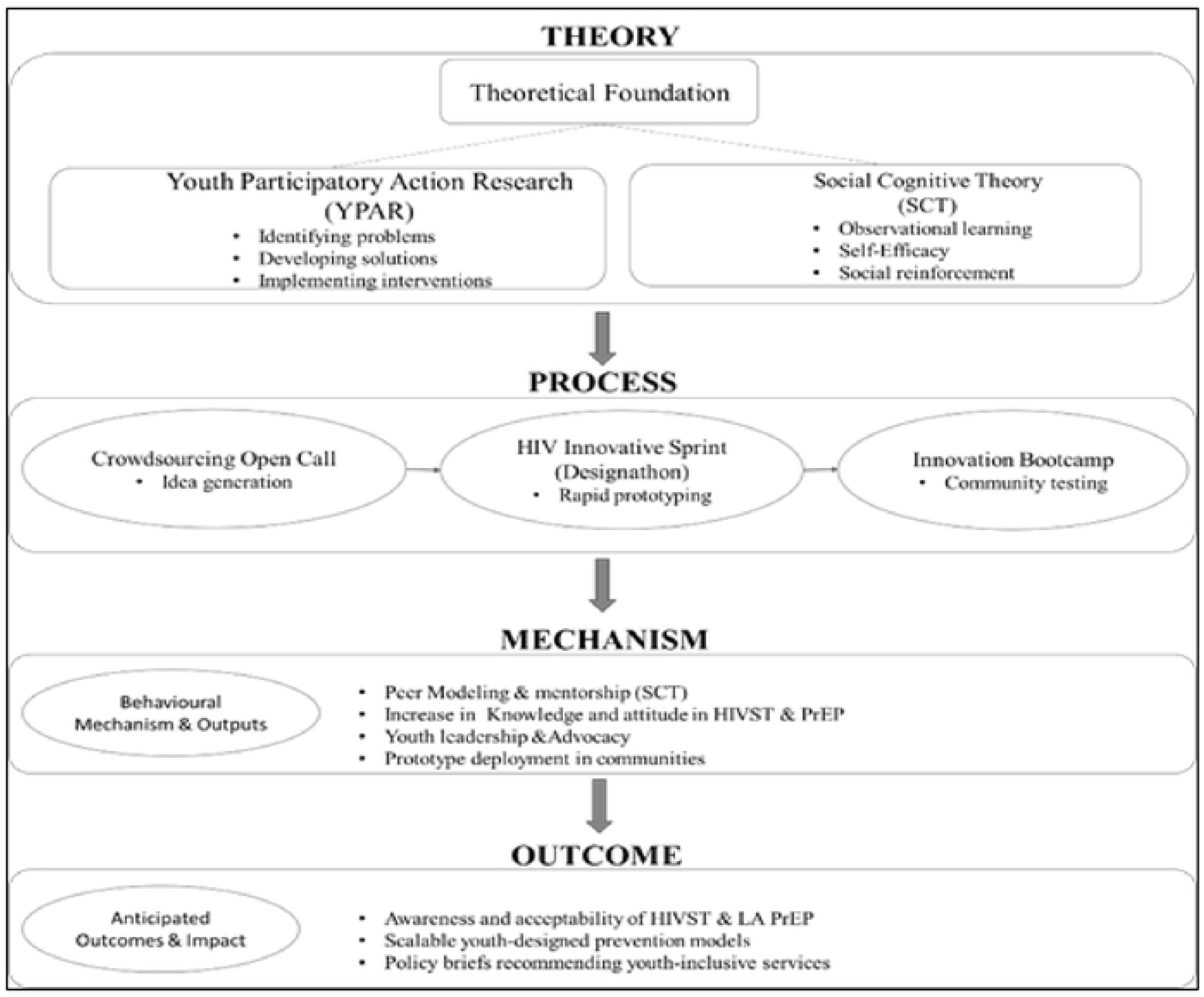
Conceptual Framework for Y-FIT Ghana Study

## Materials and Methods

### Study Objectives

The primary objective of the Y-FIT Ghana study is to develop, refine, and assess the feasibility and acceptability of youth-led strategies to increase awareness, accessibility, and uptake of HIVST, oral, and LAI PrEP among AYAs in Ghana. A secondary objective is to assess the feasibility and acceptability of these youth-generated strategies and to examine contextual factors that influence their readiness for implementation. By positioning adolescents and young adults as co-creators of HIV prevention approaches, the study aims to generate foundational evidence to inform future pilot testing, scale-up planning, and integration of youth-driven strategies into national HIV prevention efforts.

### Study Design

Y-FIT Ghana employs a participatory study design grounded in Youth Participatory Action Research (YPAR) and social innovation principles. Adolescents and young adults are engaged as active partners in knowledge generation, decision-making, and intervention design throughout the study. The study unfolds across three sequential phases: (1) a national crowdsourcing open call, (2) an HIV Innovation Sprint, and (3) an Innovation Bootcamp. Together, these phases form a structured innovation pipeline that supports youth participants in progressing from idea generation to the development of implementation-ready HIV prevention strategies. All study procedures will be conducted in accordance with the protocol approved by the Ghana Health Service Ethics Review Committee (GHS-ERC) (*ID NO: GHS-ERC-030/05/25*) and the University at Buffalo Institutional Review Board (UB IRB) *(IRB ID: STUDY00009385)*.

### Study Setting

The study will be implemented nationwide in Ghana, engaging adolescents and young adults from urban, peri-urban, and rural settings. Although recruitment will be national, core in-person activities, including the HIV Innovation Sprint and Innovation Bootcamp, will be conducted in Accra. Accra serves as a strategic hub due to its accessibility via national transportation networks, the presence of youth-serving organizations, and the availability of facilitators, mentors, and technical partners experienced in participatory innovation and youth engagement.

### Study Population and Eligibility

Participants will be Ghanaian adolescents and young adults aged 15–24 years. Eligibility will be intentionally inclusive to ensure diversity across gender identity, socioeconomic background, and geographic region. Youth may apply individually or in teams of two to four members.

Eligible participants must reside in Ghana and fall within the target age range. Prior experience in HIV prevention or research will not be required. Participants aged 18–24 will provide written informed consent. For youth aged 15–17, parental consent and participant assent will be obtained in accordance with national ethical guidelines.

### Recruitment Procedures

Recruitment will follow GHS-ERC–approved procedures and will combine digital outreach with community-based engagement. Digital recruitment will include social media campaigns on platforms commonly used by young people, including Facebook, Instagram, TikTok, WhatsApp, and X, using youth-friendly videos, graphics, and messaging that direct interested individuals to an electronic screening and application form. Complementary in-person recruitment will occur through schools, universities, vocational institutions, youth clubs, faith-based organizations, and community centers. All recruitment materials will emphasize voluntariness, confidentiality, and the opportunity to contribute to youth-led HIV prevention efforts. Study staff will review applications to confirm eligibility and to support representation across demographic characteristics and geographic regions.

### Study Status and Timeline

At the time of submission, the Y-FIT Ghana study is ongoing and has not generated outcome data related to HIV self-testing uptake, PrEP initiation, or linkage outcomes. The preparatory phase of the study, consisting of a national youth innovation open call, was conducted between May and August 2025. The open call solicited youth-generated ideas to improve youth-friendly HIV prevention strategies. This phase was designed to inform the subsequent co-creation and innovation sprint activities. Funding release delays accounted for the gap between preparatory activities in 2025 and implementation in 2026. The next phase of the study, the Innovation Sprint, is scheduled to take place between May and July 2026. During this phase, selected youth participants will engage in structured co-design sessions to refine intervention prototypes. Next will be the Bootcamp, which will take place between June and September 2026.

### Intervention Development

The Y-FIT Ghana intervention will be implemented through a three-stage social innovation pipeline designed to support youth in progressing from idea generation to the development of implementation-ready HIV prevention strategies (Figure 2). The pipeline includes a national crowdsourcing open call, an HIV Innovation Sprint, and an intensive Innovation Bootcamp. Each phase incorporates participatory design methods and youth-centered facilitation to ensure that resulting strategies reflect the preferences, priorities, and lived realities of Ghanaian adolescents and young adults. Across all phases, participants will receive structured mentorship, opportunities for peer collaboration, and guided feedback to refine prevention concepts and assess feasibility, acceptability, and implementation considerations. The pipeline is intended to prepare youth-led innovations for future small-scale testing and integration into broader HIV prevention programming.

**Figure 2:**
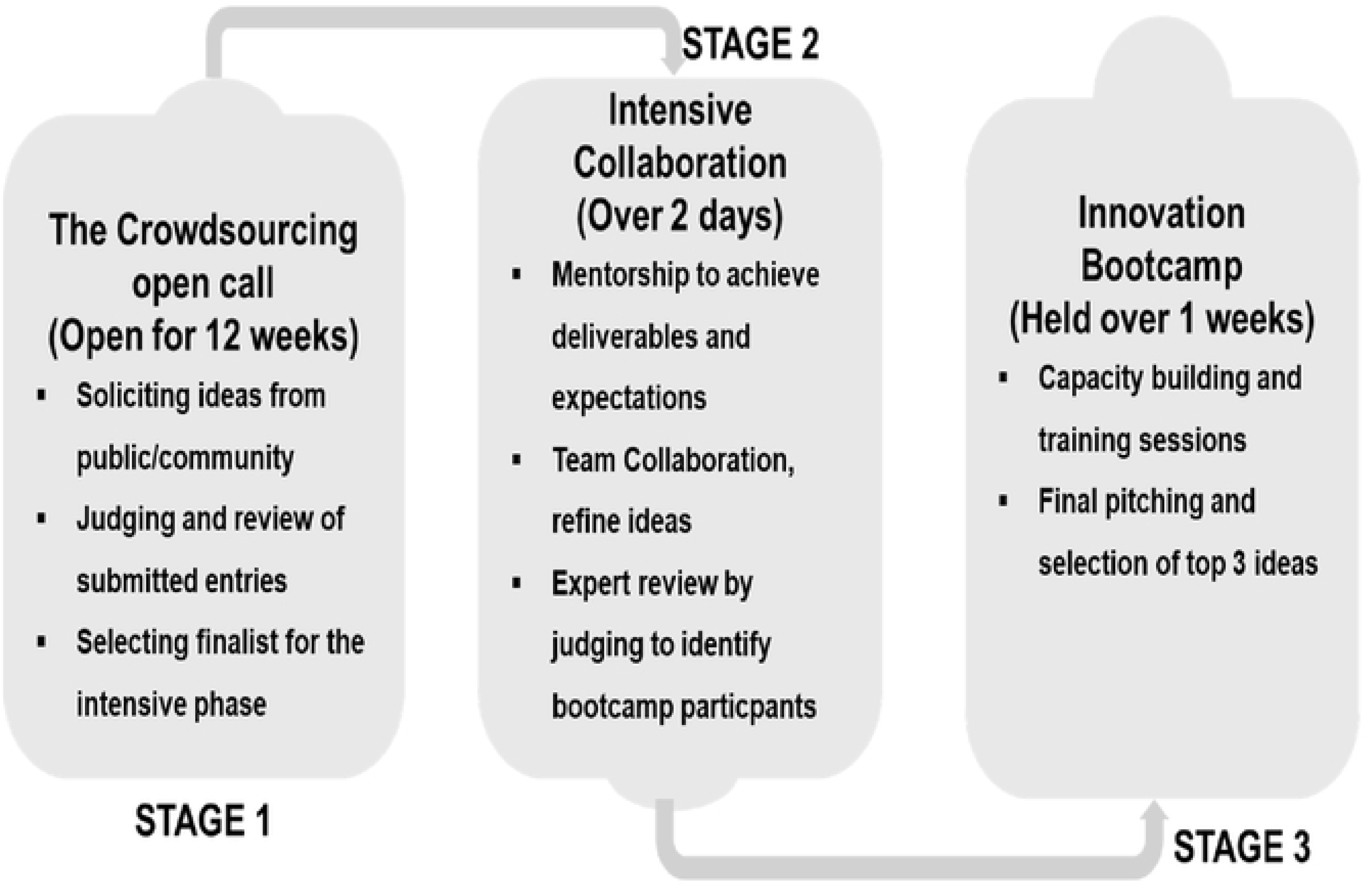
Intervention development stages

### Intervention stages

The Y-FIT Ghana intervention will be implemented through a three-stage youth-led social innovation pipeline designed to move participants from idea generation to the development of implementation-ready HIV prevention strategies (Table 1).

**Table 1.**
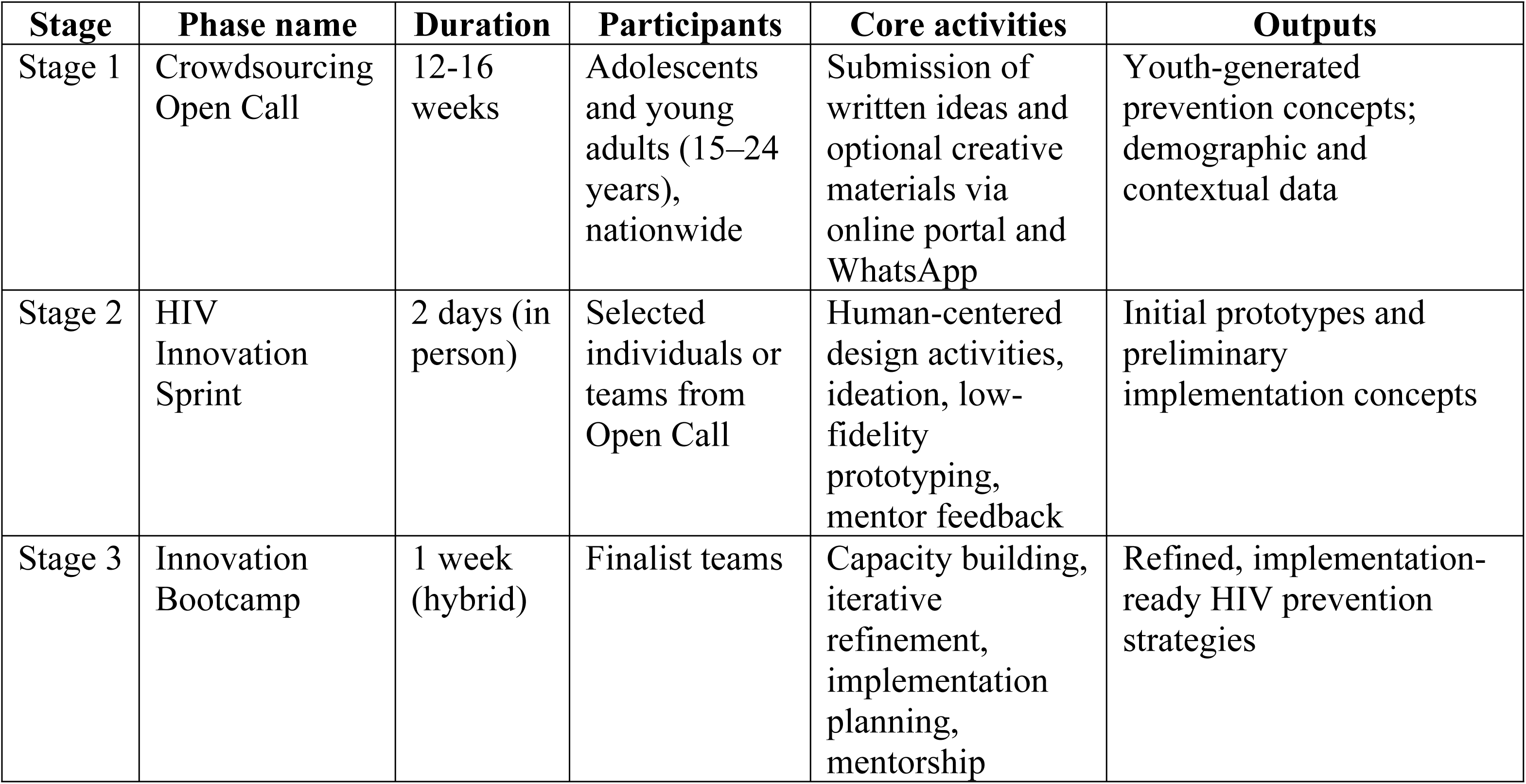
Y-FIT Ghana social innovation pipeline.

| **Stage** | **Phase name** | **Duration** | **Participants** | **Core activities** | **Outputs** |
| --- | --- | --- | --- | --- | --- |
| Stage 1 | Crowdsourcing Open Call | 12-16 weeks | Adolescents and young adults (15–24 years), nationwide | Submission of written ideas and optional creative materials via online portal and WhatsApp | Youth-generated prevention concepts; demographic and contextual data |
| Stage 2 | HIV Innovation Sprint | 2 days (in person) | Selected individuals or teams from Open Call | Human-centered design activities, ideation, low-fidelity prototyping, mentor feedback | Initial prototypes and preliminary implementation concepts |
| Stage 3 | Innovation Bootcamp | 1 week (hybrid) | Finalist teams | Capacity building, iterative refinement, implementation planning, mentorship | Refined, implementation-ready HIV prevention strategies |

#### Stage 1: Crowdsourcing Open Call

The intervention will begin with a national Crowdsourcing Open Call that will serve as the entry point for youth engagement and idea generation. Over a seven-week period, adolescents and young adults across Ghana will be invited to propose innovative strategies for increasing awareness, accessibility, and uptake of HIV self-testing and long-acting injectable PrEP among their peers. Consistent with IRB-approved procedures, the call will be widely disseminated through social media platforms, youth-serving organizations, schools, community-based institutions, and faith networks to ensure broad geographic and demographic reach. Submissions will be accepted through an online portal and WhatsApp to reduce technological barriers and enhance accessibility.

Participants will be asked to submit a 300–500-word proposal addressing two core prompts: (1) how HIV self-testing and LAI PrEP can be effectively promoted among Ghanaian youth, and (2) how young people prefer to access these prevention tools. Youth will also have the option to include a creative component—such as a poster, infographic, short video, jingle, or design sketch—to illustrate their ideas. The submission system will collect demographic information including age, gender, region, and contact details, as well as additional guardian information and consent documentation for minors aged 15–17.

All submissions will undergo eligibility screening before review by a multidisciplinary committee composed of youth leaders, HIV prevention experts, behavioral scientists, and community stakeholders. Reviewers will score entries independently using a standardized rubric evaluating innovation, feasibility within the Ghanaian context, scalability, youth-friendliness, and potential public health impact. The panel will then deliberate to identify the strongest concepts, and approximately twelve high-scoring individuals or teams will advance to the HIV Innovation Sprint. This phase will generate a diverse set of youth-driven insights and foundational concepts that will guide the structured co-creation and refinement activities in subsequent stages of the Y-FIT GH pipeline.

#### Stage 2: HIV Innovation Sprint

The second phase of the intervention will be the HIV Innovation Sprint, a focused, in-person design workshop during which semifinalist teams will refine concepts generated through the Crowdsourcing Open Call. In the literature, similar youth innovation workshops are often referred to as “designathons.” However, in the Ghanaian context, the research team elected to use the term *Innovation Sprint* due to the recent proliferation of Guinness World Record attempts — such as cook-a-thons and sing-a-thons — that generated substantial public fatigue and skepticism toward any event branded with the suffix “-thon.” The rebranding to *Innovation Sprint*, therefore, reflects both contextual sensitivity and the need to ensure that youth perceive the activity as meaningful, credible, and distinct from entertainment-based challenges dominating the national discourse.

The HIV Innovation Sprint will be held in Accra over two intensive days to facilitate sustained collaboration among youth, mentors, and facilitators trained in human-centered design and participatory research methods. Throughout the Sprint, participants will engage in structured activities designed to deepen their understanding of the problem space, clarify the needs and preferences of Ghanaian adolescents and young adults, and examine the social and structural factors influencing HIV prevention behaviors. Facilitators will lead iterative rounds of ideation, sketching, and low-fidelity prototyping to help teams translate initial ideas into concrete, youth-centered concepts.

Mentors with expertise in HIV prevention, digital communication, design, and youth development will provide continuous feedback, guiding teams as they refine their prototypes and consider the feasibility, acceptability, and cultural resonance of their emerging strategies. Teams will map user journeys, identify key touchpoints for delivering HIV self-testing and PrEP-related information, and explore delivery modalities that align with youth norms, communication channels, and trusted social networks.

By the conclusion of the Sprint, each team will have produced an initial prototype and a preliminary implementation concept describing the intended audience, delivery mechanisms, essential activities, and anticipated challenges. A review committee will evaluate these outputs for clarity, innovation, feasibility, and potential public health impact. The most promising teams will then be selected to advance to the Innovation Bootcamp, where intervention models will undergo more extensive refinement and preparation for implementation.

#### Stage 3: Innovation Bootcamp

The Innovation Bootcamp will provide an extended period of capacity building during which finalist teams will strengthen technical, conceptual, and implementation skills necessary to transform their prototypes into fully elaborated intervention models. Conducted over multiple weeks through a combination of in-person sessions and virtual mentoring, the Bootcamp will offer youth comprehensive training in effective communication, community engagement, HIV literacy, digital content creation, behavioral insights, and key concepts from implementation science relevant to feasibility, acceptability, and scalability.

Throughout the Bootcamp, youth will refine their prototypes through iterative testing, peer feedback cycles, and facilitated problem-solving sessions. Mentors will meet regularly with each team to support the development of implementation plans, guide refinement of messaging and delivery components, and help assess contextual factors that may influence uptake or sustainability. Teams will conduct small-scale field exercises, such as role-plays, informal usability testing, or brief interviews with peers, to gather early feedback and identify areas for adjustment.

By the conclusion of the Bootcamp, each team will have produced a refined prototype and an implementation-ready plan that articulates the intervention’s objectives, core components, delivery approach, target population, and monitoring indicators. The Bootcamp will culminate in a public showcase where youth present their interventions to stakeholders, technical advisors, and community representatives. Feedback from this event will inform decisions regarding which youth-led strategies will be supported during the follow-up mentorship period to prepare for pilot implementation.

### Detailed activity of the Innovation Bootcamp

The Innovation Bootcamp will be a one-week intensive capacity-building and apprenticeship program. The Bootcamp’s structure and content closely follow the procedures outlined in the approved protocol. Detailed Innovation Bootcamp activities are summarized in Table 2.

**Table 2.** Innovation Bootcamp structure and training components.

| **Activity** | **Focus area** | **Key content** | **Skills developed** |
| --- | --- | --- | --- |
| Activity 3.1 | Foundational context and user insight | HIV epidemiology, youth vulnerability, HIVST and PrEP landscape, empathy mapping, journey mapping | Problem framing, user-centered analysis |
| Activity 3.2 | Ideation and prototyping | Brainstorming, rapid prototyping, qualitative and quantitative research methods | Creative ideation, basic research literacy |
| Activity 3.3 | Implementation planning | Logic models, timelines, resource mapping, budgeting, and sustainability planning | Implementation readiness, feasibility assessment |
| Activity 3.4 | Communication and dissemination | Presentation coaching, peer review, final pitch showcase | Communication, stakeholder engagement |

### Data collection procedures

All data collection procedures will follow tools and protocols approved by the GHS-ERC (*ID NO: GHS-ERC-030/05/25*) and the UB IRB (*IRB ID: STUDY00009385*). Baseline surveys will be administered during the screening and onboarding process to capture participant demographics, HIV knowledge, and self-efficacy. During the HIV Innovation Sprint and Innovation Bootcamp, pre–post assessments will be used to evaluate changes in learning outcomes, research skills, leadership capacity, and attitudes toward HIV HIVST and PrEP. Qualitative data will be collected throughout the intervention to document participant experiences and the evolution of youth-led intervention ideas. Data sources will include semi-structured interviews, focus group discussions, participant reflection journals, facilitator field notes, observation summaries, and mentor reflections. These data will provide insight into contextual barriers and facilitators, group dynamics, and processes shaping prototype development and implementation readiness.

### Consent

The Y-FIT Ghana study has received ethical approval from the Ghana Health Service Ethics Review Committee and the University at Buffalo Institutional Review Board. All study procedures will comply with national and international ethical guidelines for research involving human participants, including provisions for voluntary participation, confidentiality, and the protection of minors. Written informed consent will be obtained from participants aged 18–24 years. For adolescents aged 15–17 years, written parental or guardian consent and youth assent will be obtained in accordance with Ghanaian ethical regulations. Consent and assent procedures will be integrated into both digital and in-person recruitment workflows to ensure participants understand the study purpose, procedures, potential risks, and anticipated benefits. All study data, including demographic information, survey responses, qualitative transcripts, screening materials, scoring rubrics, prototypes, and digital artifacts, will be stored on secure, password-protected servers accessible only to authorized study personnel. Participant identifiers will be separated from analytic datasets, and unique identification codes will be used for all analyses. Audio recordings and transcripts will be de-identified during transcription, and visual or creative submissions containing identifying information will be redacted or coded prior to analysis. Data will be managed and retained in accordance with IRB-approved data protection protocols. Study findings will be reported in aggregate, and no identifying information will be included in publications or dissemination materials. De-identified datasets may be shared through approved repositories upon request, subject to ethical and regulatory requirements.

### Data analysis

#### Overview of analytical approach

Data analysis will follow a mixed-methods strategy designed to assess feasibility, acceptability, implementation readiness, and the developmental trajectory of youth-driven HIV prevention strategies. Quantitative and qualitative data will be analyzed separately and then integrated to generate a comprehensive understanding of study outcomes. Quantitative analyses will be conducted using R, and qualitative analyses will be conducted using NVivo.

### Quantitative data management and analysis

Prior to analysis, quantitative data will undergo systematic cleaning and validation procedures, including checks for missing values, inconsistencies, and outliers. Patterns of missing data will be assessed, and appropriate methods such as complete-case analysis or multiple imputation will be applied as needed. Variables will be recoded and transformed where appropriate, and internal consistency of multi-item scales will be assessed using Cronbach’s alpha. Descriptive statistics will summarize participant characteristics, engagement patterns, and baseline measures. Pre–post changes in outcomes such as HIV knowledge, self-efficacy, leadership capacity, research skills, and attitudes toward HIVST and PrEP will be examined using paired-sample t-tests or Wilcoxon signed-rank tests, depending on distributional assumptions. For categorical outcomes, McNemar’s test will be used. Multivariable regression models will explore associations between demographic characteristics and observed changes. Prototype evaluation scores generated by mentors and judges during the Innovation Sprint and Bootcamp will also be analyzed quantitatively. Mean scores will be compared across teams, inter-rater reliability will be assessed using intraclass correlation coefficients, and associations between prototype scores and participant outcomes will be examined.

### Qualitative data management and analysis

Qualitative data will include semi-structured interviews, focus group discussions, participant reflection journals, facilitator field notes, observation summaries, mentor reflections, and youth-generated creative submissions produced during the crowdsourcing open call and subsequent intervention stages. Creative submissions may include written stories, videos, visual artwork, design sketches, songs, jingles, and other multimedia materials that reflect youth perspectives on HIVST and PrEP. All audio and video materials will be transcribed verbatim, with visual and creative content documented through detailed analytic descriptions capturing key elements such as narrative content, imagery, symbolism, tone, and intended messaging. Textual, visual, and audio data will be imported into NVivo or linked analytic memos to support systematic coding and analysis. Analysis will begin with data familiarization through repeated review of transcripts, creative materials, and field notes, accompanied by memo writing to document initial impressions and analytic insights. A hybrid inductive–deductive coding approach will be used. Deductive codes will be informed by the Consolidated Framework for Implementation Research (CFIR) to examine implementation-relevant constructs, including intervention characteristics, individual-level influences, contextual factors, and implementation processes. Inductive codes will capture emergent themes related to youth experiences, creative expression, values, social meanings, and the evolution of intervention ideas. Given the multimodal and process-oriented nature of the data, qualitative analysis will explicitly examine how youth ideas, narratives, and representations of HIV prevention evolve across intervention stages. Comparative analyses will be conducted across data sources and formats to identify shared themes, divergent perspectives, and shifts in framing from early idea generation to implementation-ready concepts.

Coding will be conducted iteratively, with regular analytic meetings to refine code definitions and resolve discrepancies. A subset of materials across data types will be double-coded to assess analytic consistency. Themes will be developed using Braun and Clarke’s six-phase approach to thematic analysis, with reflexive memos used to document analytic decisions and enhance transparency. Triangulation across interviews, observations, journals, mentor reflections, and creative submissions will be used to strengthen credibility and to examine alignment between youth-articulated priorities, observed group processes, and proposed intervention strategies. These analyses will inform assessments of feasibility, acceptability, and implementation readiness of youth-led HIV prevention approaches. Creative submissions will be analyzed as expressions of youth meaning-making and innovation rather than as artistic products, with attention to how content, form, and delivery reflect youth priorities and social contexts.

### Mixed-methods integration

Following separate quantitative and qualitative analyses, findings will be integrated using a convergent parallel mixed-methods design. Quantitative and qualitative data will be collected concurrently across intervention stages, analyzed independently, and then brought together to generate integrated interpretations related to feasibility, acceptability, and implementation readiness of youth-led HIV prevention strategies. Integration will occur at multiple points in the analytic process.

First, quantitative findings from surveys and prototype scoring, including changes in HIV knowledge, self-efficacy, leadership capacity, and attitudes toward HIVST and PrEP, will be compared with qualitative themes derived from interviews, focus groups, reflection journals, observations, mentor feedback, and creative submissions. Joint displays will be developed to visually align quantitative outcomes with qualitative explanations of how and why observed changes occurred.

Second, qualitative insights will be used to contextualize and interpret quantitative patterns. For example, improvements in knowledge or self-efficacy will be examined alongside youth narratives, creative expressions, and observational data that describe learning processes, shifts in confidence, and evolving perceptions of HIV prevention tools. Conversely, areas where quantitative change is limited will be explored qualitatively to identify contextual barriers, social dynamics, or implementation challenges.

Third, integrated findings will be used to assess the implementation potential of youth-generated intervention ideas. CFIR-informed qualitative themes will be examined alongside quantitative prototype evaluation scores to identify strategies demonstrating strong alignment with youth preferences, feasibility within the Ghanaian context, and readiness for future pilot testing. These integrated analyses will support the identification of intervention concepts with the greatest promise for reach, adoption, and sustainability. Through this mixed-methods integration, the study will generate meta-inferences that clarify not only whether youth-led innovation approaches are feasible and acceptable, but also how participatory processes and creative engagement contribute to the development of youth-friendly HIV prevention strategies.

### Ethical considerations and Declarations

This study will be performed in line with the principles of the Declaration of Helsinki. Approval will be sort from the Ethics Committee of University at Buffalo and the Ghana Health Service Ethics committee.

### Consent to participate

*Informed consent will be obtained from all individual participants 18 years and above and child assent together with parental consent obtained from participants below 18 years included in the study*.

### Dissemination Plan

Findings from the Y-FIT Ghana study will be disseminated to youth, community stakeholders, public health practitioners, policymakers, and academic audiences to support youth-centered HIV prevention efforts in Ghana. Consistent with the study’s participatory orientation, dissemination activities will prioritize accessible and equitable knowledge sharing. Study results and youth-generated outputs will be shared with participating youth and community partners through a dissemination forum in Accra and through digital channels, including social media and WhatsApp. A brief youth-friendly summary will be developed to communicate key findings and lessons learned. A technical summary of findings related to feasibility, acceptability, and implementation readiness will be shared with national and regional stakeholders, including the Ghana Health Service and Ghana AIDS Commission, to inform HIV prevention programming. Findings will also be disseminated through peer-reviewed publications and presentations at regional and international scientific conferences focused on HIV prevention, global health, and implementation science.

## Discussion

This protocol describes Y-FIT Ghana, a youth-led social innovation model designed to generate contextually grounded strategies to increase awareness and uptake of HIV self-testing (HIVST) and oral and long-acting injectable PrEP among adolescents and young adults in Ghana. By positioning youth as co-creators rather than passive recipients of HIV prevention efforts, the study departs from traditional top-down program design approaches that have often failed to resonate with young people.

A key contribution of Y-FIT Ghana is the integration of Youth Participatory Action Research and Social Innovation Theory within an implementation science–informed framework. The structured innovation pipeline supports youth progression from idea generation to early implementation planning, while remaining responsive to Ghana’s sociocultural context. The adaptation of the designathon model into an “Innovation Sprint” illustrates how participatory methods must be locally tailored to maintain credibility and engagement, particularly in youth-focused research.

The study also addresses an important gap in the introduction of emerging HIV prevention technologies. While HIVST and PrEP offer substantial promise, their uptake among young people depends on how well delivery strategies align with youth preferences, social norms, and access realities. Youth-generated outputs from Y-FIT Ghana are expected to surface communication strategies, access pathways, and peer-driven mechanisms that may be overlooked by conventional programs. By documenting feasibility, acceptability, and implementation considerations from a youth perspective, the study will generate evidence to inform future HIV prevention efforts in Ghana.

Several limitations warrant consideration. Despite broad recruitment strategies, some groups, including youth with limited digital access or those in remote settings, may be underrepresented. The study addresses this through mixed recruitment approaches and multiple submission formats. In addition, youth-led processes may privilege more confident participants; trained facilitators will use inclusive methods to support equitable participation. Finally, while youth-generated prototypes may require further refinement prior to testing, the mentorship and feedback processes embedded in the study are designed to strengthen implementation readiness. Overall, Y-FIT Ghana contributes a structured, youth-centered model for participatory innovation that balances creativity with implementation feasibility, offering insights relevant to HIV prevention programming in Ghana and similar settings.

## Conclusion

Y-FIT Ghana is expected to address a critical gap in HIV prevention research by advancing a youth-led, participatory approach to the development of HIVST and PrEP strategies. By engaging adolescents and young adults as active contributors throughout the innovation process, the study will generate contextually relevant insights to inform future pilot testing, scale-up, and integration of youth-centered HIV prevention approaches in Ghana.

## Data Availability Statement

The data generated and analyzed during this study will not be publicly shared due to ethical restrictions related to participant confidentiality, the involvement of adolescents, and the sensitive nature of HIV-related data. Data access is restricted under the approvals granted by the Ghana Health Service Ethics Review Committee and the University at Buffalo Institutional Review Board.

## Author’s Contributions

Gloria Aidoo-Frimpong led study conceptualization, design, implementation oversight, and contributed to data analysis and manuscript development. Gloria Aidoo-Frimpong & Abass Tando Abubakar drafted the original manuscript and supported study design, data curation, and analysis. Yaa Adutwumwaa Obeng, Winfred Kofi Mensa, and Daniel Selase Anyidoho contributed to data management, analysis, and manuscript review. Ernest Ortsin, Stephen Ayisi Addo, Anthony Affum Awuah, Naa Ashiley Vanderpuye, and Paul Sowah facilitated study implementation and youth engagement. Gloria Aidoo-Frimpong and Temitope Ojo provided methodological support and contributed to qualitative analysis and interpretation. Ucheoma Nwaozuru, Zhao Ni, and Juliet Iwelunmor contributed to conceptualization, interpretation, supervision, and manuscript refinement. All authors reviewed and approved the final manuscript.

## Acknowledgments

The authors acknowledge the institutional and ethical oversight provided by the University at Buffalo and the Ghana Health Service Ethics Review Committee (GHS-ERC) during the development of this study protocol. We also thank the Ghana AIDS Commission and the National AIDS Control Programme (NACP-Ghana) for their strategic guidance and technical input in aligning the study with national HIV prevention priorities.

## Y-FIT GH Team

Helen Appiah-Ampofo; Michael Gborglah; Enoch Nana Yaw Oduro Agyei; David Caleb Gborglah; Emmanuel Osei-Owusu; Robert Mawuko; Esinu Aku Adza; Senam Aku Biddah; Juvillar Fugar, Dr. Joseph Tucker, Dr. Oliver Ezechi, Dr. Chris Guure, Dr. Gamji Abu-Baare, Ms. Emma Gyamerah, Dr.Anthony Afum Adjei-Awuah, Dr. Kwasi Torpey, Perk Pomeyie, LeatherOnCall Gh, Ebenezer Asare, West Africa AIDS Foundation, Educational Assessment and Research Center, THRIVE Lab at the University at Buffalo, Belah Health Clinic, Belah Pharmacy, Lead Productions, Webrand Advertising, and StoriiLab Studios.

